# Effects of Early Career Peer Review Service on Subsequent Grant Submission Outcomes

**DOI:** 10.64898/2026.05.15.26353357

**Authors:** Adrian Vancea, Khushbu Shah Pandit, Mehmet Ertan Ornek, Dipak Bhattacharyya, Mark D. Lindner, Bruce Reed

## Abstract

Peer reviewers provide a critical service to NIH by evaluating the scientific and technical merit of grant applications. While the tangible rewards for this service are limited, many reviewers feel review service makes them better applicants, improving their grant competitiveness. However, empirical evidence for this claim is limited. This study evaluates relationships between early career peer review service and subsequent application behavior and funding outcomes.

Using NIH administrative data, applicants who served as Early Career Reviewers (ECRs) during the 2020 - 2021 council years were compared to a matched group of ECR-eligible applicants who had not served as reviewers (n=1,120 per group). To address non-random selection of ECRs, propensity score matching was used to balance groups on research field, demographics, productivity, career stage, and institutional resources. Outcomes, assessed over a three-year follow-up period, included submission volume, peer review scores, and funding outcomes for R01 and R01-equivalent applications.

ECRs submitted more applications, were more likely to have their applications discussed, and were more likely to receive a high review score than matched controls. They were also more likely to receive R01 funding. While peer review scores do not solely determine award outcomes, these findings indicate that peer review service among ECRs is associated with improved grant application outcomes.

## Introduction

The National Institutes of Health (NIH) is the largest public funder of biomedical research in the world, awarding more than $37 billion annually, mostly in the form of grants, to support research at academic institutions, medical centers, and small businesses (1). Peer review is a critical component of NIH’s highly competitive awards process. In the first tier of review, active scientists from a wide range of research institutions are recruited by scientific review officers (SROs) at the NIH Center for Scientific Review (CSR) to serve on peer review committees (study sections) to evaluate the scientific merit of grant applications. Committee evaluations of scientific merit go to the NH funding institutes’ national advisory councils which, in a second tier of review, consider them along with institute and NIH funding priorities, portfolio balance, budget, and other factors in making funding recommendations to the IC directors, who make the funding decisions (2).

Peer review service requires considerable time and uncompensated effort. Asked why they review, scientists often cite the opportunity to learn of cutting-edge ideas, and to network, in addition to altruistic motives. Reviewers also frequently say that the experience of serving as a reviewer makes them a better applicant. They believe that participating in review first-hand gives them understanding that allows them to write stronger grant applications, leading to more success in obtaining grants and pursuing their science. However, to our knowledge there is no objective, empirical evidence to support that idea.

Reviewers invited to serve on study sections are typically well established, mid-career or senior scientists with strong research records (i.e., funded grants, publications, awards, promotions, etc.). However, scientists early in their career may bring new ideas and a valuable perspective to review panels. Involving them in review is part of NIH’s commitment to developing the next generation of U.S. scientists.

A challenge for evaluating the effect of review service on performance as an applicant is that selection for review service is generally tied to indicators of scientific productivity and career advancement. That is, the characteristics that make an investigator likely to be selected as a reviewer are also those associated with strong grant application outcomes. This study addressed that confound by capitalizing on the NIH Early Career Reviewer (ECR) program (3), which enabled us to create contrasting groups of early career scientists that differed with respect to whether or not they had served as an NIH reviewer. The NIH Center for Scientific Review (CSR) created the Early Career Reviewer (ECR) program to expand and broaden the reviewer pool and provide a professional development opportunity for early career investigators. ECRs participate as reviewers in NIH study section meetings alongside experienced reviewers, gaining first-hand exposure to review criteria, scoring norms, and panel discussion processes. During the 2020 - 2021 council years, ECR eligibility required:

- Assistant Professor/equivalent at time of service
- At least 1 prior reviewed NIH application as Project Director or Principal Investigator (PD/PI)
- At least 2 first or last author peer-reviewed, research publications before service
- No prior NIH review service (except mail review)
- No prior R01 or R01-equivalent funding as PD/PI

SROs select ECRs from the eligible pool based on expertise and review needs. Additionally, SROs tend to favor those whose credentials suggest they might be able to serve as standing members in the future, making participation non-random: those recruited to serve generally have more previous grant applications, publications, and non-R01 or R01-equivalent funding than those who are not recruited. (The R01 is the predominant grant research project grant mechanisms NIH uses to support independent, investigator-initiated research.)

In this study, we examine whether early career investigators who served as ECRs perform differently as applicants compared to investigators who met ECR eligibility criteria but did not serve as reviewers. To address selection bias and confounding, we construct a carefully matched control group of ECR-eligible investigators using propensity score matching to balance the groups on key characteristics that may relate to future scientific productivity.

## Methods

### Study Design and Index Date

This is a retrospective, quasi-experimental design that uses as comparison groups ECR participants versus non-participant ECR-eligible early career scientists to test the hypothesis that NIH peer review service is associated with advantageous differences in subsequent grant submissions and outcomes. ECR service was not randomly assigned so we used propensity score matching (PSM) to create statistically matched groups of ECRs versus similar ECR-eligible investigators who did not serve. The primary estimand of interest is the average difference in outcomes between ECRs and the Control Group, corresponding to an average treatment effect (effect of ECR service) on the treated (on ECRs). The index date was the ECR’s review service date; for controls, we assigned a service-equivalent index date (see Control Group, below) and assessed outcomes during the subsequent three-year follow-up period.

### Data Sources

Reviewer service history, grant applications, peer review outcomes, award data, and institutional NIH funding were obtained from the official NIH administrative database (IMPAC II). Publication data were obtained from Dimensions via its API (link). Data extraction and integration used SQL and IMPAC II Query View Reports (QVR) interface, with processing and analyses conducted in R and Python.

### Study Population

#### ECR Group

The initial ECR cohort included 1,862 ECR-eligible investigators who were recruited and served as NIH Early Career Reviewers during the 2020 - 2021 council years (peer review meetings from September 2019 to July 2021). For the ECR group the index date was the date of review service.

#### Control Group

ECRs had to have had an application reviewed at NIH. Thus, to create a comparison cohort, we identified investigators with NIH applications during the same general period but no study section service. Starting from 240,813 investigators with NIH applications and no review service history, we applied sequential restrictions to approximate ECR eligibility. Requiring that investigators had at least one competing application reviewed in the 2020 - 2021 council years filtered the pool to about 71,000 records, restricting it to assistant professors yielded about 18,000; filtering for a single application with no R01-equivalent funding left 10,683, requiring that their application had been reviewed at CSR left 7,402 available for matching. For controls, the index date was the review meeting date of their application that was closest to October 1, 2020, the midpoint of the ECR service window. Those who were not Assistant Professor rank/equivalent, had prior NIH study section meeting service (except mail review), or who had prior R01 or R01-equivalent funding as PD/PI as of that index date, were excluded.

#### Final Analytic Sample

We applied identical final exclusions to both groups: at least two first or last-authored publications before index date, at least one previously reviewed NIH application before index date, non-missing academic age, and non-missing sex/race/ethnicity. Race and ethnicity were restricted to categories with sufficient sample sizes for stable matching (Asian, Black/African American, White; and Hispanic/Latino or Non-Hispanic). One individual with anomalously high post-index award amounts was excluded due to data quality concerns. The resulting pre-match sample included 1,504 ECRs and 4,169 controls (Fig. S1).

#### Covariates for propensity score estimation

Covariates were selected a priori to capture factors related to reviewer selection and grant outcomes. The covariates used for matching were: field of research (operationalized as study section for ECR service or application study section for controls); sex, race, and ethnicity; number of submitted applications prior to service; number of funded non-R01 or R01-equivalent applications prior to service; number of funded R01 or R01-equivalent applications prior to service (see details below); number of first- and last-authored publications prior to service; academic age (years since PhD degree, MD, MD/PhD or equivalent); and institutional NIH funding during the 2020 and 2021 council years.

For the period before the index date, R01 or R01-equivalent funding was an exclusion criterion for ECR eligibility. However, some ECRs were awarded an R01 or R01-equivalent after they were recruited for service, and they could not be dropped from serving once the application review process was underway. Those ECRs were retained in this study and receiving an R01 or R01-equivalent award in the 45-day period before the index date, the time typical for reviewer recruitment, was included as a matching criterion.

For the 3-year period after the index date, only applications submitted after the index date were counted for submission volume and funding.

### Propensity score estimation and matching

Propensity score matching (PSM) is widely used in observational studies to reduce the impact of treatment-selection bias in the estimation of treatment effects by balancing observed confounders between groups (4-7). We estimated propensity scores using logistic regression with ECR participation as the dependent variable and all matching covariates as predictors (8). Quadratic terms and two-way interactions were included for all covariates except field of research. We performed 1:1 nearest-neighbor matching without replacement using a caliper of 0.02 standard deviations of the logit of the propensity score, matching in descending order of propensity score (matchit in R). Using this approach, 1,120 ECRs (74% of the pre-match sample) were successfully matched to 1,120 controls.

### Outcomes

Outcomes were measured during the three-year period after the index date. Primary outcomes, restricted to R01 and R01-equivalent applications, were: (1) submission volume (number of NIH applications submitted), (2) number of discussed applications, (3) number of applications with high impact scores (≤ 35), (4) number of funded applications, and (5) total dollar amount awarded. For discussed applications, we used the final overall impact score (the average overall impact score of all reviewers voting in the meeting after the discussion); for non-discussed applications, we used the mean of the three assigned reviewer’s overall impact scores. All outcome analyses were also conducted including all applications (Tables S1-S4).

## Statistical analyses

Analyses were conducted on the matched samples. There is debate in the literature about whether the statistical analysis needs to account for the matched-pair nature of the data or not (4, 9-13). To confirm the validity of the results, we evaluated models that accounted for the matched-pair nature of the data (conditional logistic for binary outcomes and multilevel approaches for continuous outcomes) and models which did not account for the matched-pair nature of the data (logistic regression for binary outcomes and linear regression for continuous outcomes). The results were very similar. For simplicity we only report regression results for the models that do not consider the matched-pair nature of the data. Primary models were adjusted for the matching covariates to improve precision and to adjust for any residual imbalance in covariates, consistent with common practice in propensity score–matched analyses. Because ECR service could plausibly influence subsequent application submission behavior, we treated models additionally adjusting for submission volume as sensitivity analyses.

## Ethics and data privacy

This study used de-identified administrative data and was determined to be not human subjects research by the NIH Office of IRB Operations, with all analyses conducted under NIH data privacy and security requirements.

## Results

### Covariate Balance After Matching

We assessed covariate balance before and after matching using standardized mean differences (SMDs) and variance ratios along with other graphical diagnostics, like density plots and QQ plots (6, 8, 14). We considered SMD < 0.1 as indicative of adequate balance and variance ratios between 0.5 and 2 as acceptable (13-15). In the matched analytic sample (n=1,120 ECRs; n=1,120 controls), covariate balance improved substantially across all matching variables. Propensity score distributions were almost identical for the two matched groups (Fig. S2). After matching, standardized mean differences for all covariates were <0.1 and variance ratios fell within the prespecified acceptable range (0.5 - 2), indicating good balance between groups (Fig. S3) and adequate group comparability for outcome analysis (13-15). Table 1 presents demographics and other descriptors of the final analytic sample.

**Table 1:** Descriptive Table of the Matched Data (n’s=1,120) We report N and % for the categorical variables and the mean for the continuous variables

| Table 1: Descriptive Table of the Matched Data (n's=1,120) |  |  |
| --- | --- | --- |
| We report N and % for the categorical variables and the mean for the continuous variables |  |  |
|  | Control | ECR |
| Sex: Female | 520 (46.4%) | 529 (47.2%) |
| Race: Asian | 344 (30.7%) | 330 (29.5%) |
| Race: Black or African American | 59 (5.3%) | 58 (5.2%) |
| Race: White | 717 (64%) | 732 (65.4%) |
| Ethnicity: Hispanic | 56 (5%) | 56 (5%) |
| N of Prior Submitted Applications: Any Budget Mechanism | 5.05 | 5.06 |
| N of Prior Funded Applications: Any non-R01 or non-R01 equivalent | 0.70 | 0.71 |
| N of Prior Funded Applications: Any R01 or R01 equivalent | 0.13 | 0.14 |
| N of Prior Publications as Senior Author | 18.59 | 18.46 |
| Years Since Last Degree | 16.42 | 16.46 |
| Institution Sum of Awards Total CY 2020-2021 | \$1078000 | \$1113000 |

### Outcomes for R01 and R01-Equivalent Applications

Table 2 summarizes outcomes for R01 and R01-equivalent applications submitted during the three-year follow-up period. Compared with matched controls, ECRs submitted more applications (2.52 vs 1.40), had more discussed applications (1.41 vs 0.68), had more applications with favorable scores (≤ 35; 0.47 vs 0.20), and had more funded applications (0.43 vs 0.16) than controls (Table 2). Differences were moderate in magnitude (Cohen’s d ≈ 0.38 - 0.49; Table 2).

**Table 2:** Number of R01 and R01-equivalent Applications per Investigator: Funded, High Impact Scores, Discussed and Submitted per Investigator (n’s=1,120)

|  |  |  |  |
| --- | --- | --- | --- |
| Outcomes During 3 Years After ECR Service | ECR Mean | Control Mean | Effect Size Cohen's d |
| Number of R01 and R01-equivalent Submitted Applications | 2.52 | 1.40 | 0.45 |
| Number of R01 and R01-equivalent Discussed Applications | 1.41 | 0.68 | 0.48 |
| Number of R01 and R01-equivalent High Impact scores ($\leq 35$) | 0.47 | 0.20 | 0.38 |
| Number of R01 and R01-equivalent funded applications | 0.43 | 0.16 | 0.49 |

### Regression-Adjusted Outcomes

Regression models adjusting for matching covariates showed similar patterns (Table 3). Model 1 adjusts for the matching covariates while Model 2 also adjusts for the number of submitted applications. After additionally adjusting for R01 and R01-equivalent submission volume, differences were attenuated but remained (Table 3 – Model 2).

**Table 3:**
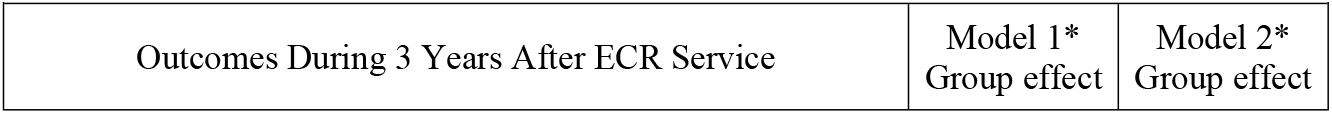
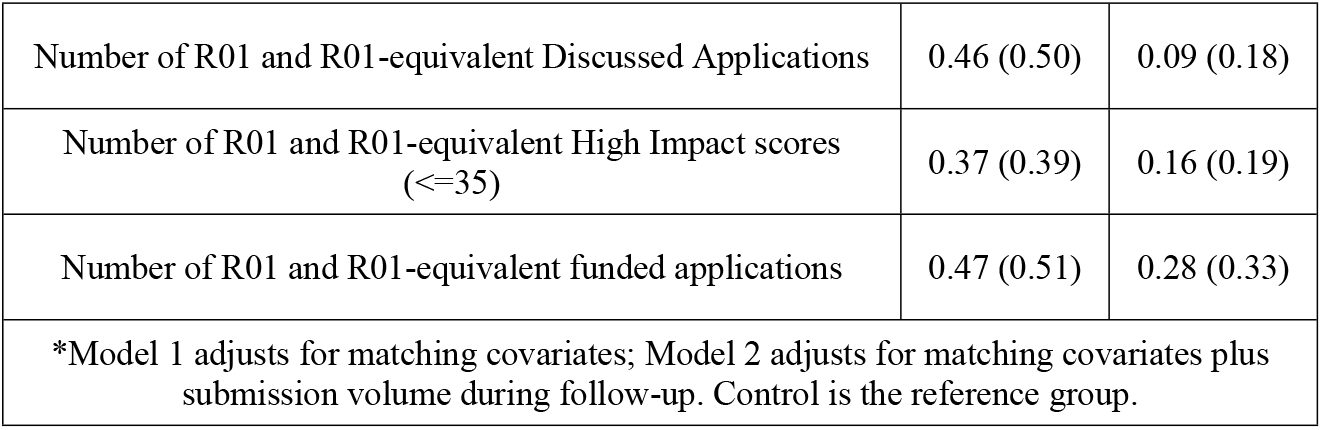
R01 and R01-equivalent applications Standardized coefficients (Cohen’s d) ECR vs Control effect from regression model

### Binary Outcomes

Logistic regression models adjusted for matching covariates (Model 1) and additionally for submission volume (Model 2). Binary outcome models showed that ECRs had substantially higher odds of achieving at least one discussed application, very favorable score (≤35) or R01 or R01-equivalent award (Table 4 – Model 1). After adjusting for R01 and R01-equivalent submission volume, ECRs remained more likely to achieve at least one favorable score (OR=2.05), and to receive at least one R01 or R01-equivalent award (OR=2.58) (Table 4 – Model 2).

**Table 4:**
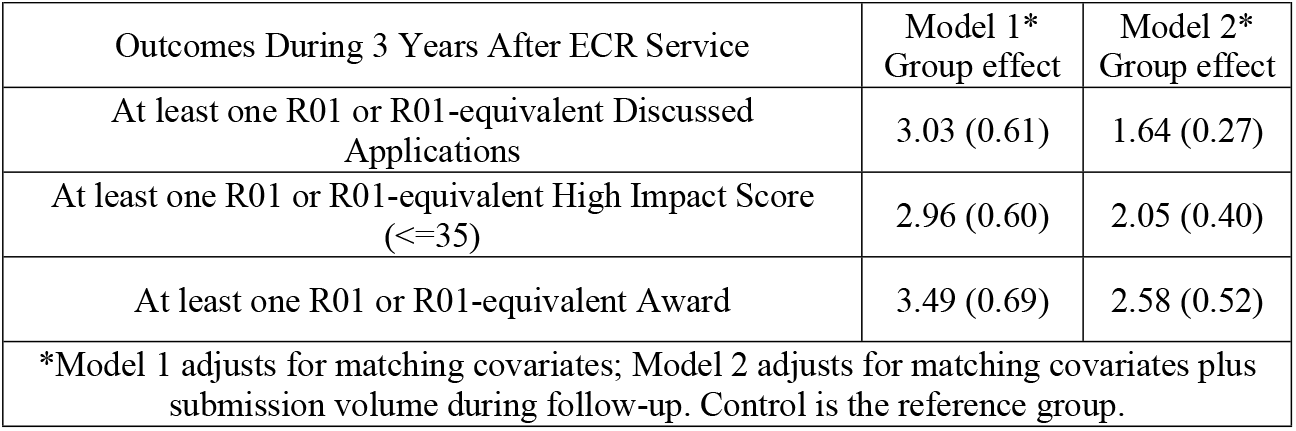
R01 and R01-equivalent applications Odds Ratios (Cohen’s d equivalent) ECR vs Control effect from regression model

|  |  |  |
| --- | --- | --- |
| Outcomes During 3 Years After ECR Service | Model 1*<br>Group effect | Model 2*<br>Group effect |
| At least one R01 or R01-equivalent Discussed Applications | 3.03 (0.61) | 1.64 (0.27) |
| At least one R01 or R01-equivalent High Impact Score (<=35) | 2.96 (0.60) | 2.05 (0.40) |
| At least one R01 or R01-equivalent Award | 3.49 (0.69) | 2.58 (0.52) |
| *Model 1 adjusts for matching covariates; Model 2 adjusts for matching covariates plus submission volume during follow-up. Control is the reference group. |  |  |

### Peer Review Scores

We also examined peer review scores as continuous outcomes. Lower scores indicate more favorable evaluations. ECRs received more favorable overall impact scores than controls for both the first and last applications submitted during follow-up (Table 5). Score differences were larger for last applications than first applications, consistent with greater improvement over time among ECRs. Mean differences were modest, and the effect sizes were small to medium.

**Table 5:** R01 and R01-equivalent Review Scores.

| Table 5: R01 and R01-equivalent Review Scores |  |  |  |
| --- | --- | --- | --- |
| Outcomes During 3 Years After ECR Service | ECR Mean | Control Mean | Effect Size<br>Cohen's d |
| First R01 or R01-equivalent Application Scores | 45.81 | 47.73 | 0.16 |
| Last R01 or R01-equivalent Application Scores | 43.52 | 47.13 | 0.29 |

## Discussion

### Findings

Early career investigators who served as NIH Early Career Reviewers (ECRs) had more favorable subsequent grant outcomes than closely matched ECR-eligible investigators who did not serve. During a three-year follow-up period, ECRs submitted more applications, were more likely to have had an application discussed, and more likely to have an application receive a high overall impact score (≤ 35). ECRs also had more funded applications and higher total award amounts. Those effect sizes were moderate. ECRs also received better peer review scores, on average, but the difference was slight and the effect size was small. The effects on binary review and funding outcomes were observed even after adjusting for submission volume. The submission-adjusted models are best interpreted as conservative sensitivity analyses: they show that group differences in outcomes are only modestly attenuated by differences in submission volume.

### Interpretation and potential mechanisms

These findings are consistent with the hypothesis that early career participation in peer review promotes more competitive subsequent grant applications. NIH shares all review criteria and reviewer training materials on public websites, and in various outreach efforts. However, direct service on a study section allows first-hand review experience. SROs train new reviewers on review policy and process, and in the meeting ECRs apply their learning, observe how more senior colleagues apply scores and review criteria, how discussions unfold, and the features that distinguish discussed and fundable applications. It is plausible that the knowledge gained contributes to writing better grant applications. This interpretation is consistent with the finding that ECR’s not only submitted more applications, but that their applications were judged to be better - more likely to be discussed, and more likely to receive a high impact score. The funding effect should be interpreted with caution. Peer review does not directly determine funding outcomes at NIH, as additional, unmeasured, factors (e.g. scientific priorities and portfolio balance) are considered in second level review and in making funding recommendations.

In addition, the mean differences in average R01 scores were small. The combination of small effects on average scores but moderate effects on likelihood of receiving a high impact score or being funded may indicate that the effects of ECR service were not evenly distributed across the group. If only minority of applicants improved substantially, their applications would be more likely to be discussed or funded (accounting for the dichotomous outcomes) but if the rest of the cohort did not improve their performance the mean score improvement would be small.

Finally, there may be other, unmeasured, advantages of study section service for early career investigators such as networking that contributes to other scientific opportunities, collaboration, or informal mentoring. Thus, these data do not prove the exact mechanism through which peer review service exerts its positive effects.

### Strengths

Strengths of this study include large samples, use of real-world data directly relevant to the NIH granting process. The matching of the “treatment” group to the control group was post-hoc, but considered multiple relevant variables and resulted in groups closely comparable on those factors. Results were consistent across multiple outcomes, all of which demonstrated positive effects with respect to important aspects of the grant submission, evaluation, and funding process.

## Limitations

Propensity score methods reduce bias from measured covariates but cannot eliminate bias from unmeasured confounding (8, 16, 17), including aspects of the ECR selection process that could not be quantified. Unknown factors that might have influenced ECR service could not be adjusted for. Administrative data are not collected primarily for evaluation; measurement error may exist for some covariates (e.g., publication linkage, institutional characteristics) and could affect matching and estimates. The three-year follow-up captures an important early-career window but does not address longer-term trajectories. Finally, inferences apply most directly to ECRs and comparable controls and may not generalize to all reviewers, and all investigators (8, 13, 14).

## Conclusion

Early career scientists who served in NIH peer review through the ECR program subsequently submitted more R01 applications. Although their mean review score were only slightly better, ECRs had more applications discussed, received more top review scores, had more applications funded, and received more NIH funding than did propensity score matched counterparts. Differences in numbers of applications submitted did not substantially attenuate the other differences observed. While causal inference is limited by the observational design and potential unmeasured confounding, the consistency of findings across multiple outcomes and analytic approaches is consistent with the frequently heard self-report of reviewers that serving in NIH peer review made them a better grant applicant.

## Data Availability

A copy of the data set is uploaded with this submission.

## Acknowledgements

The views expressed in this manuscript are those of the authors and do not necessarily reflect the official policy or position of the Department of Health and Human Services, the National Institutes of Health, or the United States Government. Publication counts were based on metadata as of 06/10/2025 from the Dimensions platform provided by Digital Science (www.dimensions.ai). Access was granted under license agreement with the NIH Center for Scientific Review.

## Figure Captions

**Fig. S1:**
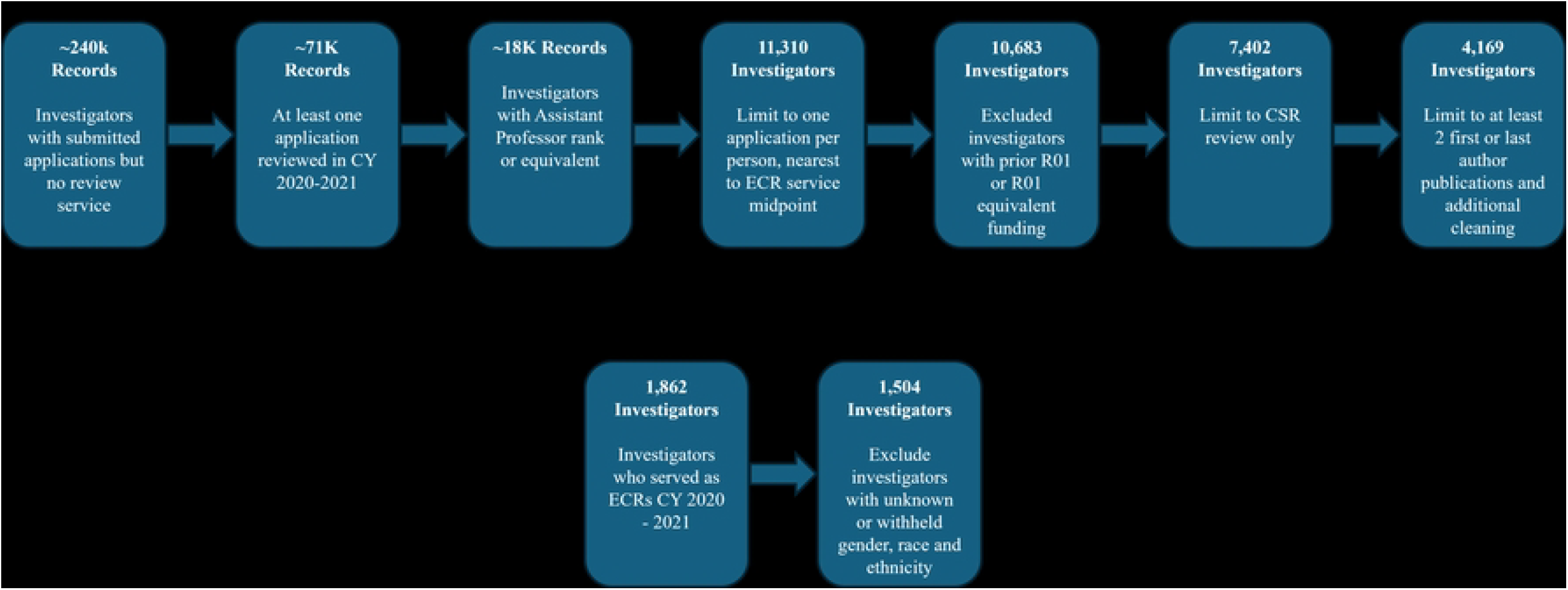
Control Group and ECR Group Sample Size. Inclusion/exclusion criteria and their effect on the sample size within each of the comparison groups.

**Fig. S2:**
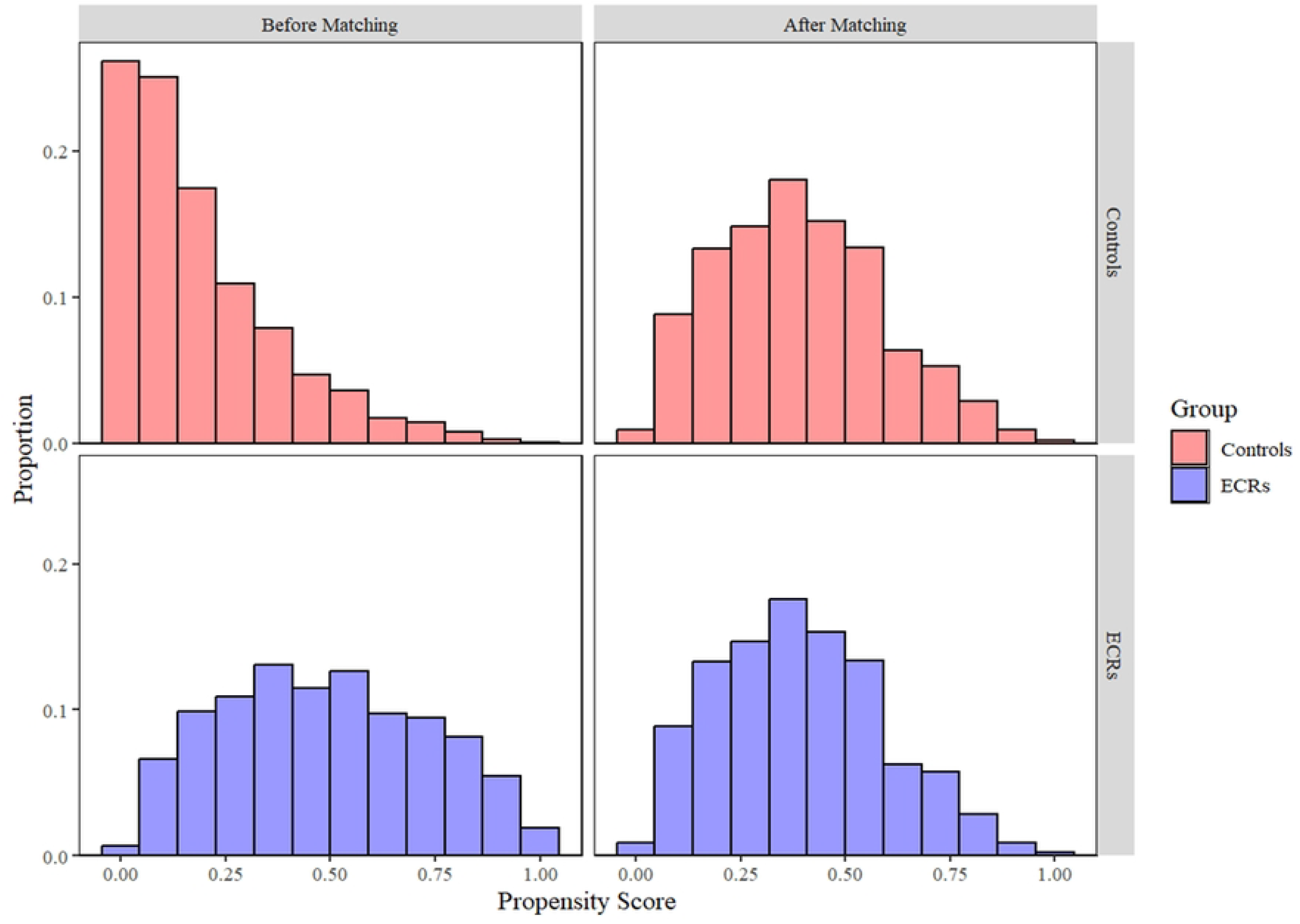
Distribution of propensity scores before and after matching. Top: Control group. Bottom: ECR group.

**Fig. S3:**
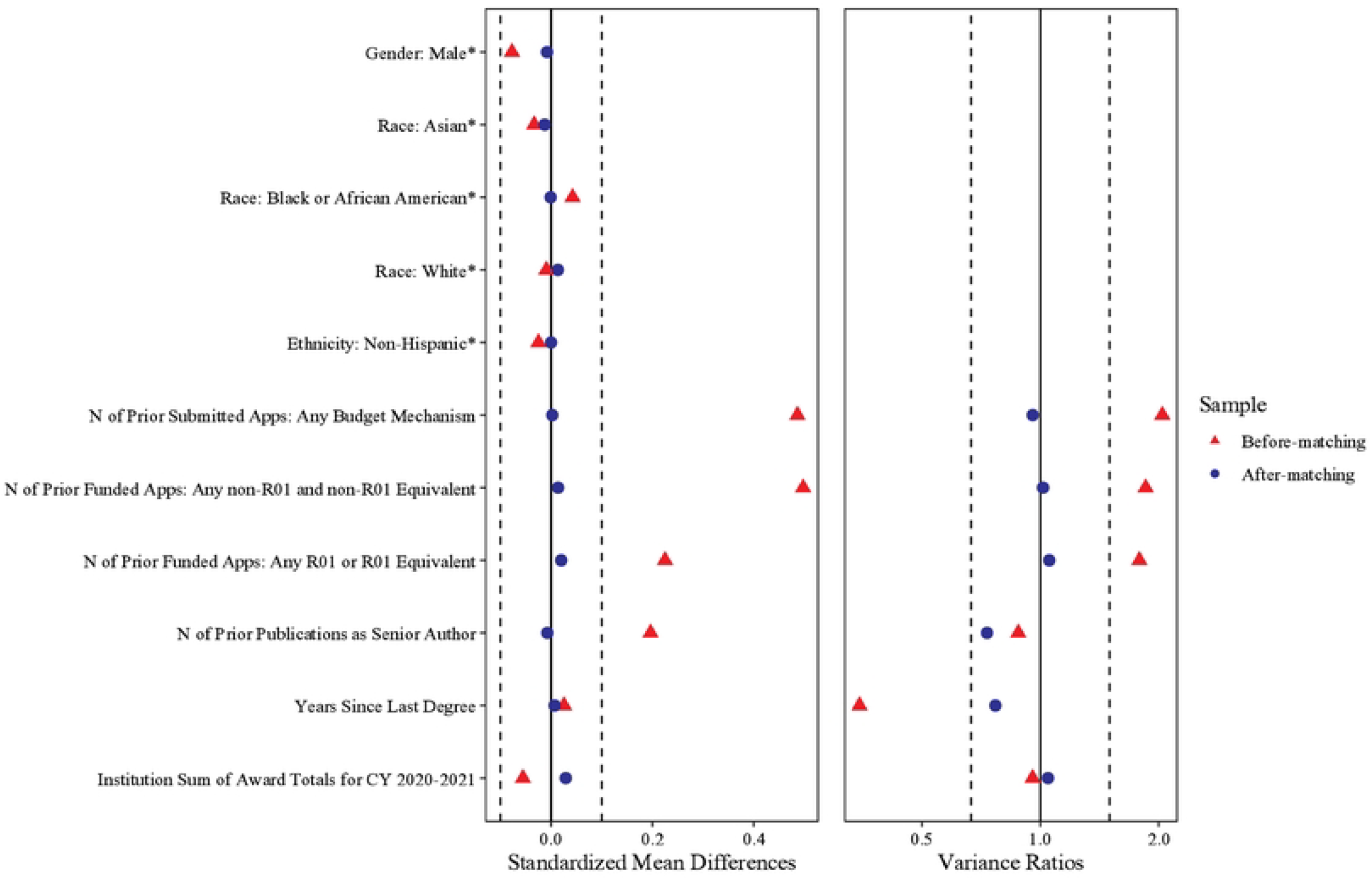
Covariate balance: Left: standardized mean differences. Right: variance ratios. Red symbols for before matching (n=4,169 Controls vs n=1,504 ECRs); Blue is data after matching (n=1,120 vs n=1,120)

**Table S1: All Applications Outcomes per Investigator:** Number of Submitted, High Impact Scores, Discussed, and Funded Applications; Total $ Awarded per Investigator (n’s=1,120)

**Table S2: All applications: Standardized coefficients (Cohen’s d) ECR vs Control effect from regression model**

**Table S3: All applications: Odds Ratios (Cohen’s d equivalent) ECR vs Control effect from regression model**

**Table S4: All Applications Review Scores**

